# Anti-phospholipid antibodies in COVID-19 are different from those detectable in the anti-phospholipid syndrome

**DOI:** 10.1101/2020.06.17.20134114

**Authors:** Maria Orietta Borghi, Asmaa Beltagy, Emirena Garrafa, Daniele Curreli, Germana Cecchini, Caterina Bodio, Claudia Grossi, Simonetta Blengino, Angela Tincani, Franco Franceschini, Laura Andreoli, Maria Grazia Lazzaroni, Silvia Piantoni, Stefania Masneri, Francesca Crisafulli, Duilio Brugnoni, Maria Lorenza Muiesan, Massimo Salvetti, Gianfranco Parati, Erminio Torresani, Michael Mahler, Francesca Heilbron, Francesca Pregnolato, Martino Pengo, Francesco Tedesco, Nicola Pozzi, Pier Luigi Meroni

## Abstract

**Background:** Critically ill patients with coronavirus disease 2019 (COVID-19) have a profound hypercoagulable state and often develop coagulopathy which leads to organ failure and death. Because of a prolonged activated partial-thromboplastin time (aPTT), a relationship with anti-phospholipid antibodies (aPL) has been proposed, but results are controversial. Functional assays for aPL (i.e., lupus anticoagulant) can be influenced by concomitant anticoagulation and/or high levels of C reactive protein. The presence of anti-cardiolipin (aCL), anti-beta2-glycoprotein I (anti-β_2_GPI) and anti-phosphatidylserine/prothrombin (aPS/PT) antibodies was not investigated systematically. Epitope specificity of anti-β_2_GPI antibodies was not reported.

**Objective:** To evaluate the prevalence and the clinical association of aPL in a large cohort of COVID-19 patients, and to characterize the epitope specificity of anti-β_2_GPI antibodies.

**Methods:** ELISA and chemiluminescence assays were used to test 122 sera of patients suffering from severe COVID-19. Of them, 16 displayed major thrombotic events.

**Results:** Anti-β_2_GPI IgG/IgA/IgM were the most frequent in 15.6/6.6/9.0% of patients, while aCL IgG/IgM were detected in 5.7/6.6% by ELISA. Comparable values were found by chemiluminescence. aPS/PT IgG/IgM were detectable in 2.5 and 9.8% by ELISA. No association between thrombosis and aPL was found. Reactivity against domain 1 and 4-5 of β_2_GPI was limited to 3/58 (5.2%) tested sera for each domain and did not correlate with aCL/anti-β_2_GPI nor with thrombosis.

**Conclusions:** aPL show a low prevalence in COVID-19 patients and are not associated with major thrombotic events. aPL in COVID-19 patients are mainly directed against β_2_GPI but display an epitope specificity different from antibodies in antiphospholipid syndrome.

## 1. Introduction

Critically ill patients with coronavirus disease 2019 (COVID-19) have a profound hypercoagulable state and often develop thrombosis in veins, arteries and in the microcirculation [1,2]. Recent analysis showed several coagulation abnormalities in these patients, including prominent elevation of fibrin/fibrinogen degradation products (i.e., D-dimer) and a prolonged activated partial-thromboplastin time (aPTT). While high levels of D-dimer are consistent with sustained activation of the clotting and fibrinolytic cascades, the combination of prolonged aPTT and both arterial and venous thrombosis was, however, surprising, and it is reminiscent of a clinical scenario known as antiphospholipid syndrome (APS) [3].

Looking at the causes for aPTT prolongation, recent studies have shown that lupus anticoagulant (LA) can be detected in a significant percentage of COVID-19 samples [4-6]. Since LA is often caused by aPL, these findings support the idea that aPL may play a role in COVID-19[7]. However, it is important to point out that LA is a very sensitive assay and its outcome can be influenced by several factors, most notably heparin administration [8] and a profound inflammatory state characterized by high levels of C reactive protein (CRP) [9,10]. Both of them are present in COVID-19 patients [11].

Another method to detect aPL that is in principle insensitive to anticoagulation and other confounding agents relies on the detection and quantification of autoantibodies using solid phase assays[3]. Using this method, the presence of aPL was recently reported in a handful of case reports and small cohorts of patients [4,6,7,12,13]. While encouraging, this data is limited and its interpretation remains controversial, with some investigators proposing an important role of aPL in COVID-19 patients[7] while others suggesting a very poor correlation between aPL and thrombotic events [14]. There is no information on the antigen specificity of COVID-19 aPL in comparison with APS antibodies. Such information and a larger study, possibly multicenter, may be instrumental to clarify the real clinical value of these autoantibodies.

## 2. Materials and methods

### 2.1 Patients

A total of 122 patients were enrolled from two COVID-19 referral centers in Lombardia. All patients tested positive to SARS-CoV-2, and classified as severe or critical COVID-19 [11]. Mean age was 68.5 (± SD 16.4) years; 77 were men and 45 women. No diagnosis of previous autoimmune diseases was made; six patients had a thrombotic event (three arterial and three venous) in the past clinical history. Eighty-seven patients suffering from APS were also tested for anti-cardiolipin (aCL) and anti-β_2_GPI IgG/IgM [15].The study was approved by the Ethics Committees (Istituto Auxologico Italiano 3-04-2020 − Milan and ASST Spedali Civili NP4187 - Brescia).

### 2.2 Detection of aPL

Anti-cardiolipin and anti-β_2_GPI IgG/IgA/IgM were detected by chemiluminescence immunoassay (CIA; Quanta Flash, Inova, San Diego, CA, US) and a home-made ELISA as described [15,16]. Anti-β_2_GPI domain 1 IgG (anti-D1) were detected by CIA[15,16], IgG anti-D4-5 by a home-made ELISA, as described [15,16]. Anti-phosphatidylserine/prothrombin (aPS/PT) IgG/IgM were detected by a commercial ELISA as reported[17].

### 2.3 Statistical analysis

Data were analyzed using R v3.4.0. Descriptive statistics was used to summarize data. Associations and differences between categorical or continuous variables were tested by Fisher’s exact test and non-parametric Mann-Whitney test, respectively. A p-value <0.05 was considered statistically significant.

## 3. Results

### 3.1 Patients

Table 1 reports the median with minimum and maximum values for different coagulation and inflammation parameters in 122 severe or critical COVID-19 patients. In particular, prolonged aPTT (>30 sec) was found in 57.6% while PT INR values were above the cut-off in 24.8% of the cases. Most of the patients (120/122) were on anticoagulation with low molecular weight heparin (70% on therapeutic and the remaining on prophylactic dosage). Despite anticoagulation, we observed sixteen thrombotic events (13.1%, 8 in veins and 8 in arteries). These statistics are in agreement with previous reports[2,18-23] and document a systemic inflammation and a coagulopathy in our patients.

**Table 1.** Coagulation and inflammation parameters expressed as median with minimum and maximum in severe or critical COVID-19 patients.

|  | D-dimer<br>μg/L | CRP<br>mg/dl | Ferritin<br>μg/L | IL-6<br>ng/L | White cells<br>n/μl | Neutrophils<br>n/μl | Platelets<br>n x 10<sup>3</sup>/μl | PT<br>ratio | aPTT<br>sec | Fibrinogen<br>mg/dl |
| --- | --- | --- | --- | --- | --- | --- | --- | --- | --- | --- |
| COVID-19 | 984.47 | 126.99 | 1024 | 25.1 | 8000 | 6600 | 350 | 1.196 | 30.13 | 521 |
|  | 200-40234 | 0.1-470.3 | 55-9002 | 3-496 | 2500-12900 | 1560-12510 | 60-800 | 0.9-6.9 | 21-75.4 | 202-840 |
| Normal range | < 500 | 0.00-0.05 | 30-400 | < 10 | 4300-10500 | 1800-8100 | 140-450 | ≤ 1.2 | < 30 sec | 200-400 |

### 3.2 Anti-cardiolipin and anti-β_2_GPI antibody testing

In the APS field, testing for LA is not recommended when patients are on heparin, since the presence of heparin, even if neutralized, may lead to false positive results [8]. Likewise, high levels CRP, such as those found in our cohort of patients, have been shown to prolong aPTT independently from the presence of aPL[9,10]. On these bases, the presence of aPL was researched using solid phase assays, and not LA. First, we investigated the presence of aCL and anti-β_2_GPI, two APS classification criteria[3]. Testing was independently performed in Milan and Brescia, using harmonized methodologies[24]. The prevalence of COVID-19 patients positive for aCL and anti-β_2_GPI IgG/IgA/IgM detected by ELISA and CIA is summarized in Table 2. The ELISA raw data are shown in Fig. 1. We found IgG/IgM aCL in 5.7/6.6% of patients, whereas anti-β_2_GPI IgG/IgA/IgM were found in 15.6/6.6/9.0% of patients. Similar values were obtained for aCL antibodies using CIA (Table 1), whereas a slightly lower sensitivity was obtained for anti-β_2_GPI antibodies [25]. The positivity for aCL and anti-β_2_GPI antibodies was at medium/low titer in contrast with the medium/high titers found in the control group of primary APS (Figure 1). There is no association between aPL positivity and thrombotic events.

**Table 2.** Prevalence of COVID-19 patients positive for aPL.

|  | ELISA |  | CIA |  |
| --- | --- | --- | --- | --- |
| | aCL | a$\beta_2$GPI | aCL CIA | a$\beta_2$GPI CIA |
| IgG | 5.7 (7)* | 15.6 (19) | 9.8 (12) | 5.0 (6) |
| IgM | 6.6 (8) | 9.0 (11) | 6.6 (8) | 5.0 (6) |
| IgA | nd | 6.6 (8) | 2.5 (3) | 0.8 (1) |

\*Values are expressed as percentage (n) of positive patients. aCL: anti-cardiolipin antibodies;
a $\beta_2$ GPI: anti- $\beta_2$ glycoprotein I antibodies; ELISA: enzyme linked immunosorbent assay; CIA:
chemiluminescence immunoassay; nd: not done.

**Figure 1.**
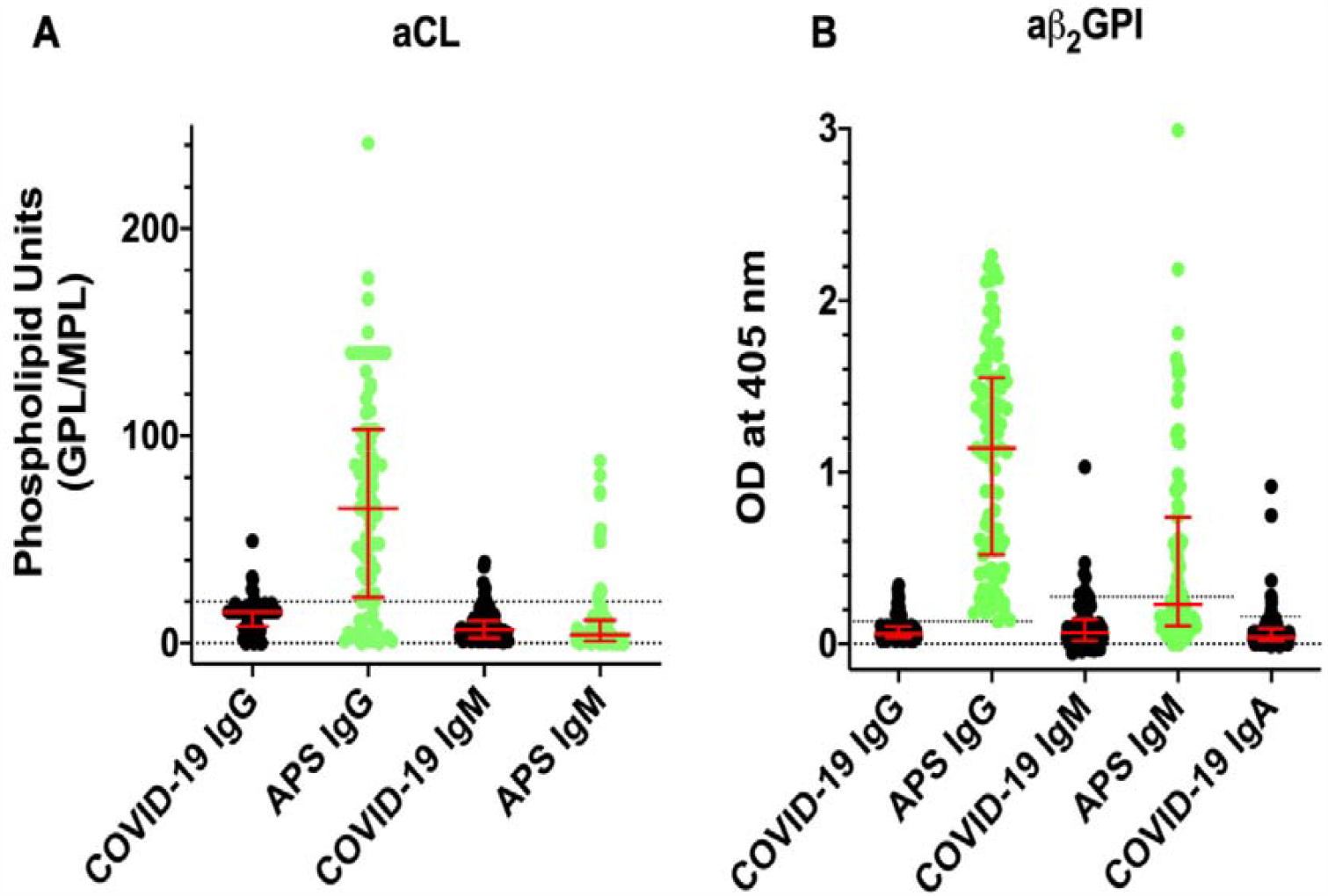
Titers of aCL and aβ_2_GPI antibodies detected by ELISA in COVID-19 patients (black, n=122) and comparison with APS patients (green, n=86). Values are expressed as median levels [first and third quartile]. **Panel A:** aCL. From the left to the right: COVID-19 IgG: 15 [8 − 15]; APS IgG: 65 [22 – 103]; COVID-19 IgM: 6.2 [2.6 − 10.8]; and APS IgM: 4.0 [1 − 11]. **Panel B**: aβ_2_GPI. From the left to the right: COVID-19 IgG: 0.06 [0.04 – 0.10]; APS IgG: 1.14 [0.52– 1.55]; COVID-19 IgM: 0.065 [0.02 – 0.142]; APS IgM: 0.23 [0.105 – 0.741]; and COVID-19 IgA: 0.04 [0.02 – 0.09]. Cutoff values are aCL IgG/IgM 20 phospholipid units (GPL/MPL); aβ_2_GPI IgG/IgM/ IgA ELISA 0.13, 0.27 and 0.16 optical units (OD), respectively.

### 3.3 Epitope characterization of anti-β_2_GPI antibodies

Fifty-eight sera were also tested with D1 and D4-5-coated plates in order to characterize their epitope specificity. Fig. 2B shows that three out of 58 samples reacted with D1, while in Fig. 2C, three samples tested positive for D4-5. None of the sera was positive for both domains and all displayed a weak reactivity with no association with thrombosis.

**Figure 2.**
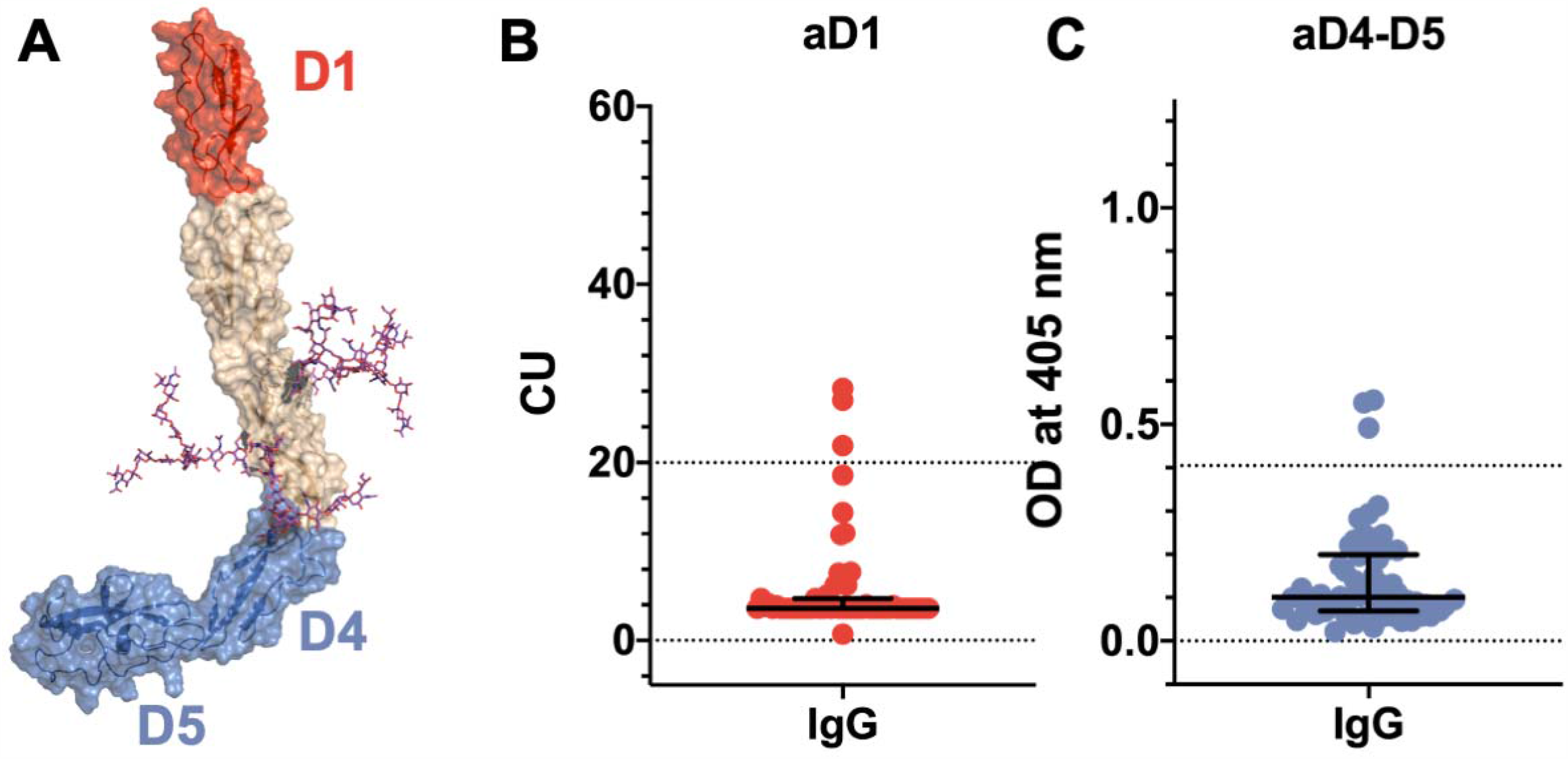
Epitope specificity of anti-β_2_GPI antibodies in COVID-19 patients. **Panel A**. Three-dimensional structure of β_2_GPI solved at 2.4 Å by X-ray crystallography (PDI ID: 6V06 [32]) displaying the positioning of the fragments used in this study. The N-terminal D1 is shown in red. The C-terminal D4-5 fragment is shown in blue. N-linked glycosylations are shown as magenta stick. Titers of anti-D1 **(panel B)** and anti-D4-5 antibodies **(panel C)** in 58 COVID-19 patients detected by chemiluminescence and ELISA, respectively. Values are expressed as median levels [first and third quartile]. Anti-D1(aD1): 3.6 [3.6 – 4.7]. Anti-D4-D5 (aD4-D5): 0.10 [0.068 – 0.199]. Cutoff values are >20 chemiluminescent units (CU) and >0.405 optical units (OD) for aD1 and aD4-D5, respectively.

### 3.4 Anti-phosphatidylserine/prothrombin antibody testing

Prolonged aPTT (>30 sec) was found in 57.6% of the patients. Although aPS/PT are not included in the APS classification laboratory tests, they can be associated with a prolonged aPTT and with the presence of LA[17]. Consequently, we looked at the presence of aPS/PT antibodies in our cohort and we found fifteen out of 122 sera positive for aPS/PT (12.3%), mostly of the IgM isotype (12 out 15) and at a low titer (Fig. 3). There was no association between prolonged aPTT and the presence of aPS/PT antibodies nor with thrombotic events in our COVID-19 cohort.

**Figure 3.**
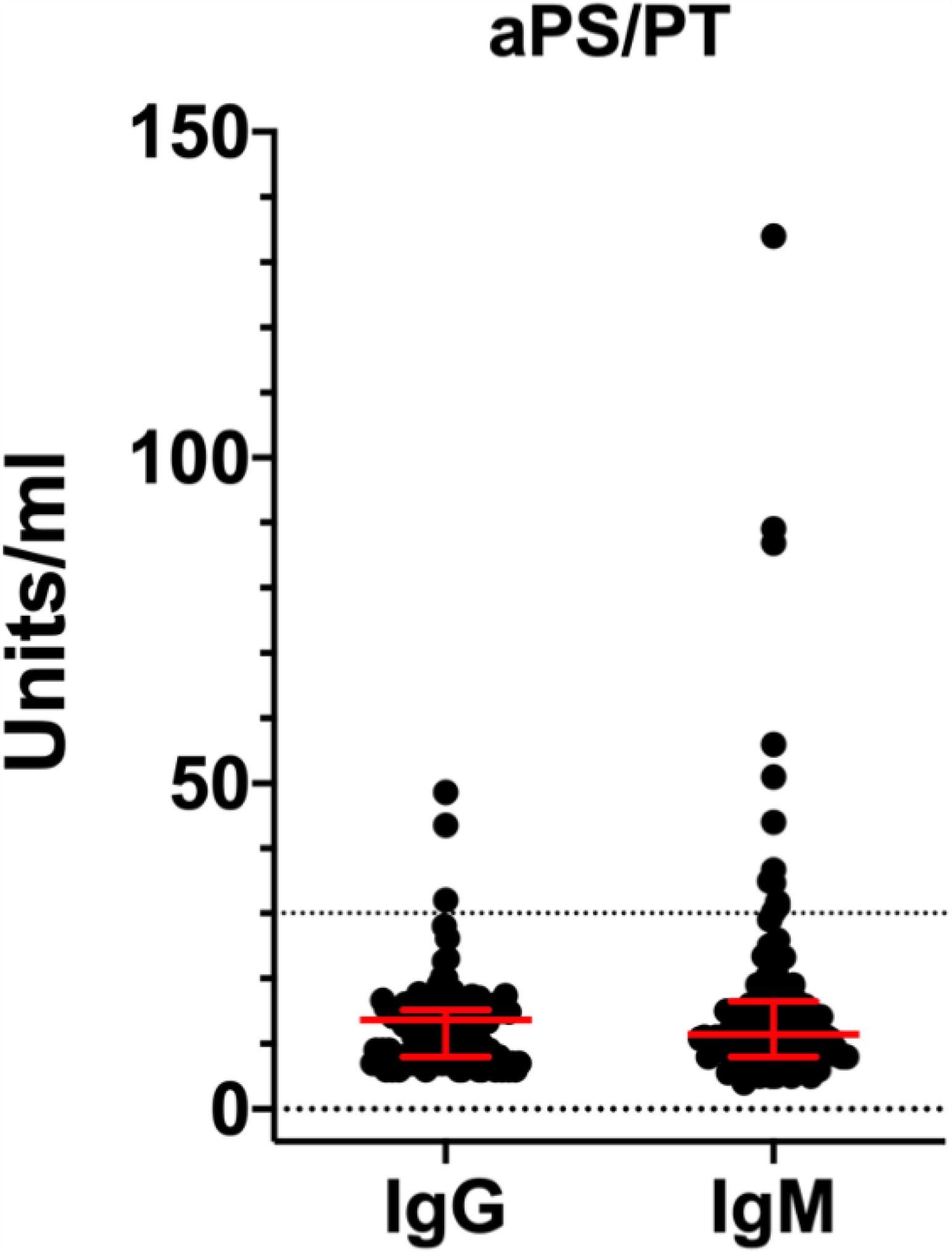
Titers of aPS/PT antibodies detected by ELISA in COVID-19 patients. Values are expressed as median levels [first and third quartile]. aPS/PT IgG:13.6 [8 to 15.2]; aPS/PT IgM:11.4 [8 to 16.5] IgM. Cut-off of the assays was 30 units/ml.

## 4. Discussion

Taken together, our data shows a low prevalence of classification criteria aPL in COVID-19 patients. In this regard, our study confirms recent studies obtained with smaller cohorts of patients[4,14,23]. Importantly, our data also shows that aPL are slightly more reactive towards β_2_GPI-coated plates as compared to CL-coated ones and that, regardless of the nature of aPL, there is no association between aPL positivity and thrombotic events (p=1).

A striking difference between the autoantibody profile in COVID-19 patients as compared to the one in APS concerned the titers of aPL. Medium/low aPL titers were consistently found in the patients with COVID-19. By contrast, medium/high titers are usually found in APS patients (Fig. 1). This difference suggests that aPL found in COVID-19 may be different from aPL found in APS and led us to further investigate the epitope specificity of anti-β_2_GPI antibodies. We focused on autoantibodies directed against the N-terminal domain 1 (anti-D1) or the C-terminal domains 4-5 (anti-D4-5) of the molecule[16] (Fig. 2A). This is because anti-D1 antibodies are associated with an increased risk of thrombosis and pregnancy complications in APS[15,16,26]. By contrast, anti D4-5 antibodies are associated neither with vascular nor obstetric APS manifestations[15,27]. Furthermore, anti D4-5 antibodies are also reported at high levels in the so called asymptomatic aPL carriers and are frequently found in non-APS (e.g. patients with leprosy, atopic dermatitis, atherosclerosis and in children born to mothers with systemic autoimmune diseases) [27]. We found that three out of 58 samples reacted with D1, and three samples tested positive for D4-5. None of the sera was positive for both domains and all displayed a weak reactivity. Although the number of the investigated sera is relatively small, this finding is quite different from the results found in APS in which almost all the sera positive for the whole β_2_GPI molecule also reacted with domain D1 at high titer[15,26]. Furthermore, at variance with APS patients, none of the anti-D1 positive patients displayed thrombotic events[26].

Approximately fifty-seven per cent of COVID-19 patients has prolonged aPTT. Yet, only a small proportion of COVID-19 patients carry aCL and anti-β_2_GPI antibodies. This suggests that other factors must be responsible for the prolonged aPTT phenomenon. Since aPS/PT can be associated with a prolonged aPTT and with the presence of LA[17], we tested our cohort for aPS/PT antibodies. We found a small percentage (12.3%) of positive sera, mostly of the IgM isotype (12 out 15) and at a low titer. Again there was no association between prolonged aPTT and the presence of aPS/PT antibodies nor with thrombotic events in our COVID-19 cohort. This indicates that aPS/PT are not responsible for the prolongation of aPTT nor are predictors of adverse clinical outcomes. Furthermore, in contrast to what we would have expected in APS[28], we found no associations between the presence of aPS/PT, aCL and anti-β_2_GPI antibodies. This data is in line with the unusual epitope specificity of anti-β_2_GPI antibodies documented in Fig. 2, supporting the hypothesis that aPL found in COVID-19 patients are different from aPL found in APS patients. Whether COVID-19 aPL are similar to the ones found in other infectious diseases such as HCV, HBV and HIV[29] remains to be determined.

In conclusion, while medium/high aPL titers with D1 specificity are associated with vascular events in APS, low antibody titers with reactivity against β_2_GPI epitope(s) different from D1 or D4,5 can be found in COVID-19. This may explain the lack of association with thrombotic events in COVID-19. In addition, our data do not support the hypothesis that aPL can be the main cause of prolonged aPTT in these patients. Although low titer aPL are not predictive of vascular events in the APS, it is important to keep in mind that COVID-19 patients suffer from an acute form of systemic inflammation with complement activation[30], which may be responsible for endothelial perturbation. In this context, since β_2_GPI can accumulate on the activated endothelium at high density, even low titers of aPL may become pathogenic thus potentiating or even triggering thrombus formation, especially when anticoagulation is suspended. A comparable condition in which low titers of aPL can cause substantial damage is seen in obstetric APS, where high levels of β_2_GPI can be found in the placenta[31]. Hence, while transitory aPL are likely to be clinically irrelevant in COVID-19 patients as in other infections[29], detection of aPL may be useful for identifying patients potentially at risk of thrombosis after the hospital discharge. Accordingly, an anticoagulant prophylaxis could be justified before a confirmatory assay[3].

## Data Availability

Data are available at the corresponding authors

## 5. Authorship

M.O. Borghi, M. Pengo, A. Tincani, F. Franceschini, F. Tedesco, N. Pozzi, P.L. Meroni designed the study; S. Blengino, G. Parati, F. Heilbron, M. Pengo, M.G. Lazzaroni, M.L. Muiesan, M. Salvetti collected clinical samples; E. Garrafa, D. Curreli, G. Cecchini, C. Bodio, C. Grossi, S. Piantoni, S. Masneri, F. Crisafulli, D. Brugnoni, E. Torresani, M. Mahler, L. Andreoli, performed research; M.O. Borghi, A. Beltagy, F. Pregnolato, F. Tedesco, N. Pozzi, P.L. Meroni analyzed data; M.O. Borghi, F. Tedesco, N. Pozzi, P.L. Meroni wrote the manuscript. All authors reviewed and approved the manuscript.

## 6. Acknowledgment

The study was in part supported by IRCCS Istituto Auxologico Italiano - Ricerca Corrente 2019 (PL Meroni), a grant from the Italian Ministry of Foreign Affairs and International Cooperation (MAECI) for foreign citizens and Italian citizens living abroad (A. Beltagy) and a National Institutes of Health Research Grant HL150146 (N. Pozzi). The Authors would like to thank: Drs. N. Carabellese and G. Martini (Department of Laboratory Diagnostics; ASST Spedali Civili, Brescia, Italy) for their valuable collaboration; all the physicians of the COVID-19 Units of the IRCCS Istituto Auxologico Italiano (Milan) and the ASST Spedali Civili (Brescia).

